# Mortality and Sudden death risk in middle-aged persons with epilepsy — a UK Biobank study

**DOI:** 10.1101/2023.07.26.23293226

**Authors:** Ravi A. Shah, C. Anwar A. Chahal, Shaheryar Ranjha, Ghaith Sharaf Dabbagh, Babken Asatryan, Ivan Limongelli, Mohammed Khanji, Fabrizio Ricci, Federica De Paoli, Susanna Zucca, Martin Tristani-Firouzi, Erik K. St. Louis, Elson L. So, Virend K. Somers

## Abstract

**Background:** Sudden death is the leading cause of mortality in medically refractory cases of epilepsy. Younger persons with epilepsy (PWE), particularly those <40 years, have higher all-cause mortality than those without. However, data are conflicting about mortality and burden of cardiovascular disease (CVD) in middle-aged PWE.

**Objective:** Determine all-cause and sudden death-specific mortality and burden of CVD in PWE in a middle-aged population.

**Methods:** Using UK Biobank, we identified 7,786 (1.6%) participants with a diagnosis of epilepsy; 566 individuals with prior history of stroke were excluded. The 7,220 PWE comprised the study cohort with the remaining 494,676 without epilepsy as the comparator group. PWE were identified based on clinical diagnostic code (validated) or self-reported diagnosis at assessment centre interview. Prevalence of CVD was determined using validated diagnostic codes. Cox proportional hazards regression were used to assess all-cause mortality and sudden death risk, in PWE vs those without epilepsy.

**Results:** Hypertension, coronary artery disease, heart failure, valvular heart disease, and congenital heart disease were all more prevalent in PWE. Arrhythmias including atrial fibrillation/flutter (12.2% vs 6.9%; p<0.01), bradyarrhythmias (7.7% vs 3.5%; p<0.01), conduction defects (6.1% vs 2.6%; p<0.01), and ventricular arrhythmias (2.3% vs 1.0%; p<0.01), as well as cardiac implantable electric devices (4.6% vs 2.0%; p<0.01) were all more common in PWE compared to comparator group. PWE had higher all-cause mortality (HR 3.9 [95% CI, 3.01-3.39]), higher sudden death-specific mortality (HR 6.65 [95% CI, 4.53-9.77]) both adjusted for age, sex and comorbidities; and were almost 2 years younger at death [68.1 vs 69.8; p<0.001].

**Conclusions:** PWE have markedly higher burden of CVD including arrhythmias and heart failure. Middle-aged PWE have increased all-cause and sudden death specific mortality and higher burden of CVD. While efforts have focused on SUDEP in the young, further work is required to elucidate mechanisms underlying all-cause mortality and sudden death risk in PWE of middle age, to identify prognostic biomarkers and develop preventative therapies in PWE. **Keywords**: Sudden Death, Epilepsy, Cardiovascular disease, UK Biobank, Epidemiology

**Clinical Perspective:** *What is new?:*

- This is the large prospective cohort study of middle-aged (40-69 years) PWE, reporting a high prevalence of CVD, including hypertension, coronary artery disease, heart failure, valvular heart disease, congenital, atrial fibrillation and ventricular tachycardia.
- PWE had a consistent 3-fold higher all-cause mortality and 6-fold higher sudden death-specific mortality than matched controls, even after multivariable adjustment.
- Of known SUDEP risk factors, only male sex and higher resting heart rates were associated with increased mortality in persons with epilepsy, but intellectual disability and polypharmacy were not (which have been reported in younger patients as risk factors).

*What are the clinical implications?:*

- Evaluation of PWE who are middle-aged should include screening for CV disease.
- This work highlights an excess burden of CVD and mortality amongst middle-aged persons with epilepsy, requiring research to identify mortality mechanisms so that this can translate to improve outcomes.

## 1. Introduction

Epilepsy affects 1-2% of the general population^1^ with significant morbidity and mortality. Younger persons with epilepsy (PWE), particularly those less than 40-years-old, have significantly higher all-cause mortality.^2^ Sudden unexpected death in epilepsy (SUDEP) is the leading mode of death in medically refractory epilepsy and has become a public health priority for policymakers.^3^ It is estimated that 17% of all deaths caused by epilepsy are SUDEP and its incidence is estimated between 0.33-6.3 per 1000-person years.^2, 4^ The definition of SUDEP is “sudden, unexpected, witnessed or unwitnessed, nontraumatic and nondrowning death in patients with epilepsy, with or without evidence for a seizure and excluding documented status epilepticus, in which postmortem examination does not reveal a toxicologic or anatomic cause of death”.^5^ Investigating the incidence and mechanisms of SUDEP is challenging as most cases are unwitnessed and systematic evaluation, including postmortem examination is rarely performed.^6^ in design.

Identified risk factors for SUDEP include age (18-40 years), male sex, intellectual disability, high frequency of generalized tonic-clonic seizures (GTCS), nocturnal GTCS, duration of epilepsy, antiepileptic polytherapy, sleep disordered breathing and QT prolongation.^7–10^ Many SUDEP events occur at night, including with patients found dead in the prone position, and face down in the pillow, similar to sudden infant death syndrome (SIDS).^10–13^ This raises potential overlap between SUDEP and SIDS and may indicate cardiac arrhythmias as potential mechanisms. Prior studies were mostly observational, retrospective, had small sample sizes, and susceptible to Berkson’s bias.^2, 14–17^

Data on middle-aged PWE are lacking. In this study, we aimed to leverage the UK Biobank (UKBB) to investigate prevalence of cardiovascular disease (CVD), all-cause and sudden death-specific mortality in a large population of middle-aged PWE. Further, we sought to investigate the potential factors associated with increased mortality or sudden death in PWE.

## 2. Materials and Methods

### 2.1 Study Population

The study population was derived from the UKBB study, a prospective cohort of more than 500,000 individuals aged 40-69 years recruited from across the UK. In brief, between 2006 and 2010, participants were recruited and completed a touchscreen questionnaire, were interviewed, and underwent anthropometric measurements, vital sign assessments, and blood sampling. Information on baseline characteristics, self-reported medical conditions and medications, and measurements, including electrocardiograms (ECG) was obtained. The UKBB has access to external data sources, which provide information, in the form of ICD-10 diagnostic codes and OPCS-4 operation codes, from hospital admissions and primary care records. Access to national death and cancer registries allows for information on time and cause of death, including death certificate data. Follow-up data are available until February 2021.

### 2.2 Procedures

A cohort of PWE was identified by electronic health records (ICD-10 diagnostic codes of G40.0-9, G41.0-9, and F80.3) or self-reported epilepsy on the enrollment questionnaire (field 20002 in the UKBB) (**eTable 1**). Patients who develop seizures following stroke are now considered to have epilepsy; however, we excluded these patients as stroke is a confounder for mortality and our focus was all-cause mortality and sudden death risk assessment in primary epilepsy. In this cohort study, PWE in the UKBB, who met eligibility criteria, comprised the epilepsy cohort (cases). The remainder of the UKBB participants was used as a referent comparator group (comparator-group). A mixture of ICD diagnostic codes, OPCS4 codes and self-reported conditions and operations were used to determine the comorbidities and causes of death of interest. Epilepsy subtypes were defined as per the International League of Epilepsy.^18^ Rhythm abnormalities, heart failure and coronary artery disease were defined based on prior publications.^19–21^ Sudden death was defined by ICD10 code of I46.1 and R96.0 or sudden death, unexpected death, unascertained cause of death, cardiac arrest, or ventricular fibrillation found within the “Description of cause of death” field (40010). Anti-epileptic medications were defined by the list of antiepileptic medications from the Medicines and Healthcare products Regulatory Agency (MHRA). Antiarrhythmic medications were classified based on Lei *et al*.^22^ A full list of codes and data fields used for phenotype characterization can be found in **eTable 1**.

This study was approved by the Mayo Clinic Institutional Review Board. Access to UKBB was provided under application 48286. This study complies with the Declaration of Helsinki and the work is covered by the ethical approval for UKBB studies from the NHS National Research Ethics Service on 17th June 2011 (Ref 11/NW/0382) and extended on 18 June 2021 (Ref 21/NW/0157). Written informed consent was obtained from all participants; participants who withdrew consent were excluded. The reporting of this cohort study follows the STROBE statement (**eTable 2**). Corresponding authors had full access to all the data in the study and take responsibility for its integrity and the data analysis. A through search for studies reporting relevant findings has been done to compare our results with already published data (**eTable 3**). Patients or the public were not involved in the design, conduct, reporting, nor dissemination plans of our research.

### 2.4 Statistical analysis

In **Supplemental methods**

## 3. Results

### 3.1 Baseline characteristics

Of 502,462 UKBB participants, 7,786 (1.6%) were identified as PWE; of these 566 were excluded due to a prior history of stroke preceding the diagnosis of epilepsy. Patients who have epilepsy and subsequently develop stroke/TIA were included. The remaining 494,676 without epilepsy were designated as a comparator group **(Figure 1**).

**Figure 1.**
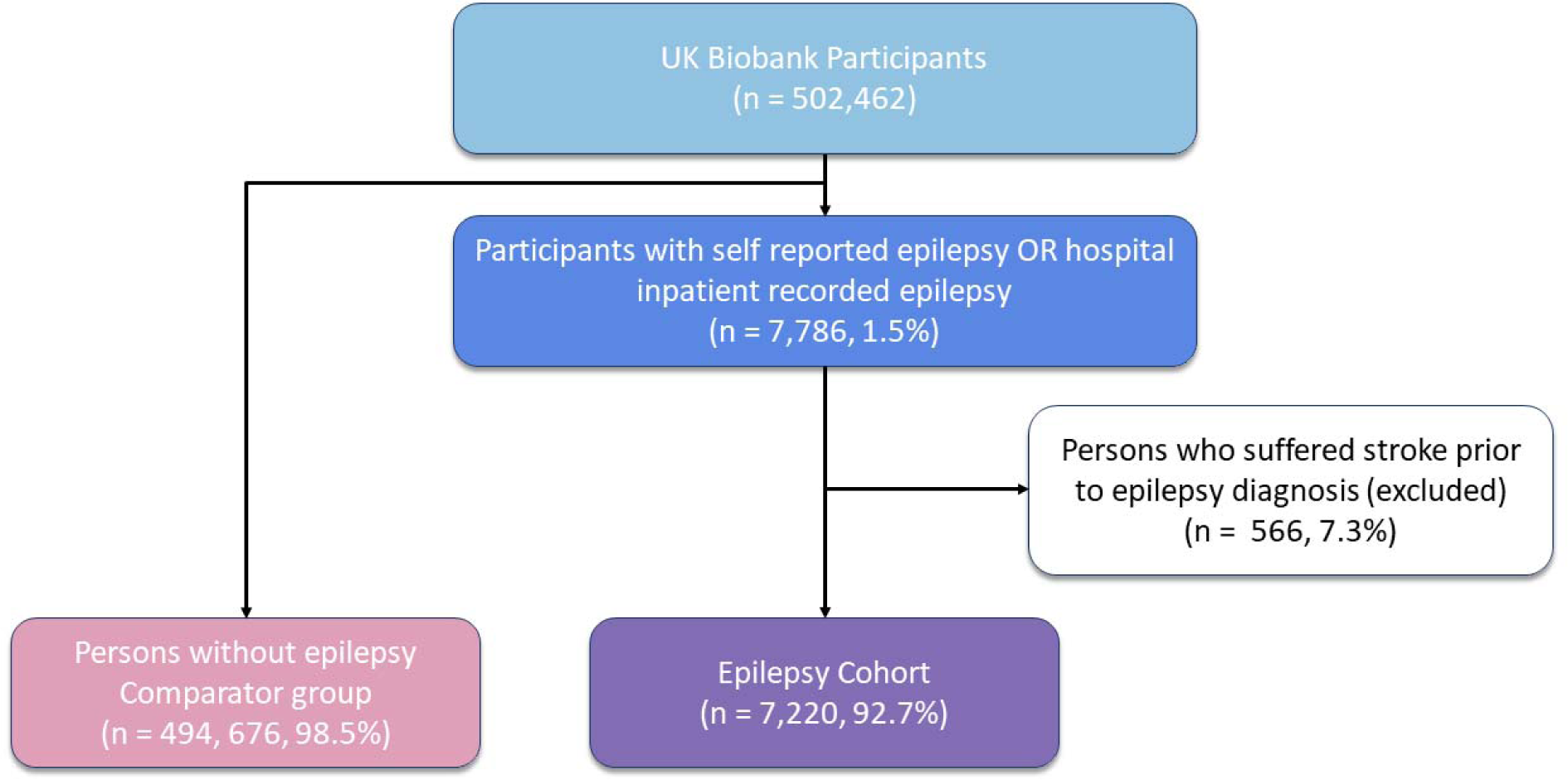
Selection of study cohorts Flowchart demonstrating the process of selecting the epilepsy cohort and comparator group from the entire UK Biobank population.

Baseline characteristics are shown in **Table 1**. There were significantly more females among the comparator cohort than PWE [49.9% vs 54.5%; p<0.001]. PWE and controls were at similar age at recruitment but had slightly greater BMI compared to controls [28.0 kg/m2 vs 27.4 kg/m2; p<0.001].

**Table 1.**
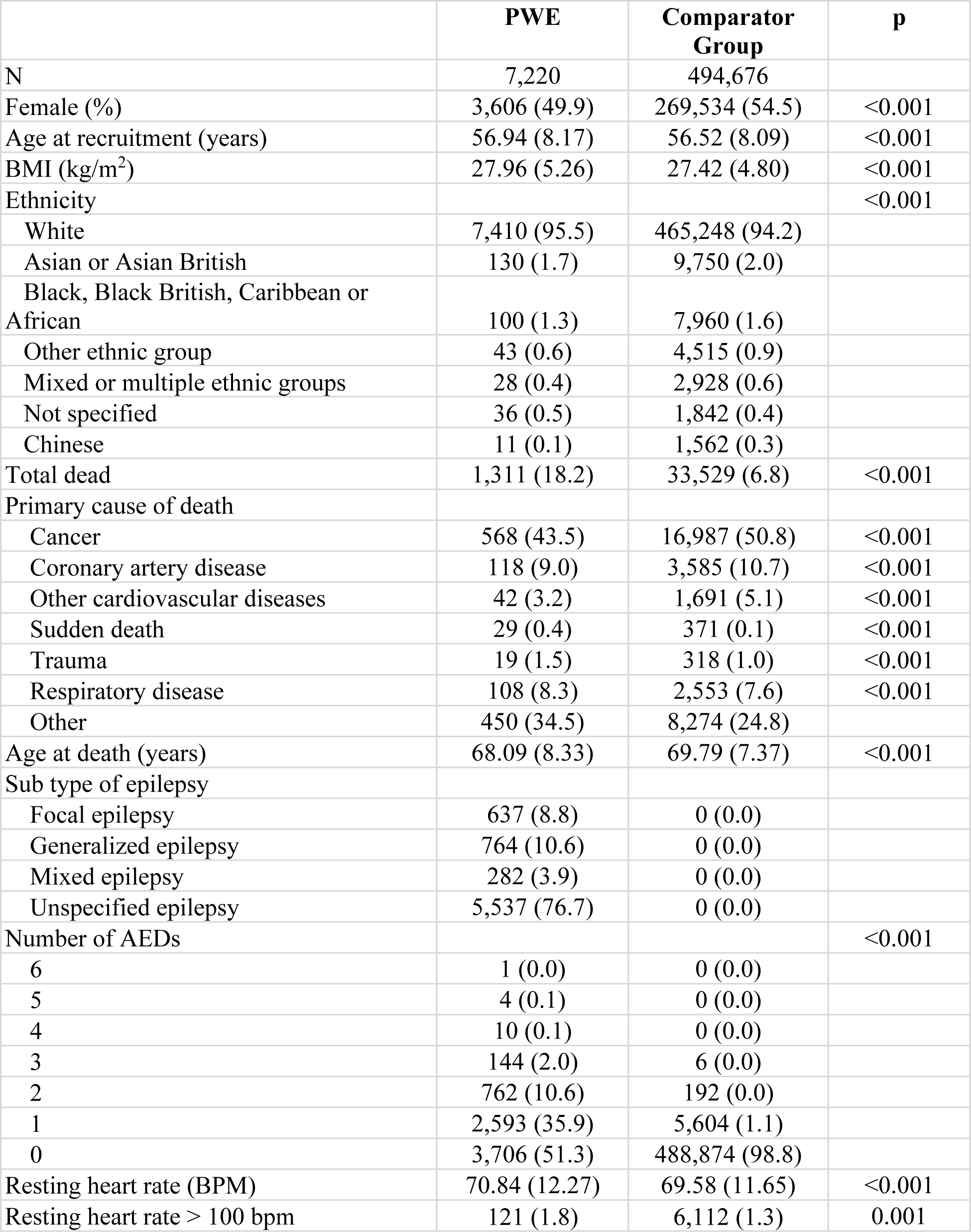
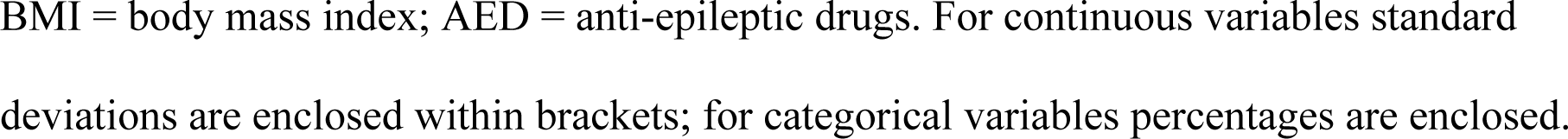
Baseline characteristics of PWE and comparator cohorts.

|  | **PWE** | **Comparator Group** | **p** |
| --- | --- | --- | --- |
| N | 7,220 | 494,676 |  |
| Female (%) | 3,606 (49.9) | 269,534 (54.5) | <0.001 |
| Age at recruitment (years) | 56.94 (8.17) | 56.52 (8.09) | <0.001 |
| BMI (kg/m<sup>2</sup>) | 27.96 (5.26) | 27.42 (4.80) | <0.001 |
| Ethnicity |  |  | <0.001 |
| White | 7,410 (95.5) | 465,248 (94.2) |  |
| Asian or Asian British | 130 (1.7) | 9,750 (2.0) |  |
| Black, Black British, Caribbean or African | 100 (1.3) | 7,960 (1.6) |  |
| Other ethnic group | 43 (0.6) | 4,515 (0.9) |  |
| Mixed or multiple ethnic groups | 28 (0.4) | 2,928 (0.6) |  |
| Not specified | 36 (0.5) | 1,842 (0.4) |  |
| Chinese | 11 (0.1) | 1,562 (0.3) |  |
| Total dead | 1,311 (18.2) | 33,529 (6.8) | <0.001 |
| Primary cause of death |  |  |  |
| Cancer | 568 (43.5) | 16,987 (50.8) | <0.001 |
| Coronary artery disease | 118 (9.0) | 3,585 (10.7) | <0.001 |
| Other cardiovascular diseases | 42 (3.2) | 1,691 (5.1) | <0.001 |
| Sudden death | 29 (0.4) | 371 (0.1) | <0.001 |
| Trauma | 19 (1.5) | 318 (1.0) | <0.001 |
| Respiratory disease | 108 (8.3) | 2,553 (7.6) | <0.001 |
| Other | 450 (34.5) | 8,274 (24.8) |  |
| Age at death (years) | 68.09 (8.33) | 69.79 (7.37) | <0.001 |
| Sub type of epilepsy |  |  |  |
| Focal epilepsy | 637 (8.8) | 0 (0.0) |  |
| Generalized epilepsy | 764 (10.6) | 0 (0.0) |  |
| Mixed epilepsy | 282 (3.9) | 0 (0.0) |  |
| Unspecified epilepsy | 5,537 (76.7) | 0 (0.0) |  |
| Number of AEDs |  |  | <0.001 |
| 6 | 1 (0.0) | 0 (0.0) |  |
| 5 | 4 (0.1) | 0 (0.0) |  |
| 4 | 10 (0.1) | 0 (0.0) |  |
| 3 | 144 (2.0) | 6 (0.0) |  |
| 2 | 762 (10.6) | 192 (0.0) |  |
| 1 | 2,593 (35.9) | 5,604 (1.1) |  |
| 0 | 3,706 (51.3) | 488,874 (98.8) |  |
| Resting heart rate (BPM) | 70.84 (12.27) | 69.58 (11.65) | <0.001 |
| Resting heart rate > 100 bpm | 121 (1.8) | 6,112 (1.3) | 0.001 |

BMI = body mass index; AED = anti-epileptic drugs. For continuous variables standard deviations are enclosed within brackets; for categorical variables percentages are enclosed.

Classification into subtypes of epilepsy was available in 1,683 PWE (23.3%). Of these, 45.5% had generalized epilepsy, 37.8% focal epilepsy, and 16.7% mixed epilepsy. PWE were more likely to be taking anti-epileptic medication than controls (48.7% on at least 1 AED vs. 1.2% controls). A subset of 100 of the comparator group on anti-epileptics were reviewed to determine the indications for these medications; indications included chronic pain, neuropathic pain and bipolar disease; none of the controls was taking AED for epilepsy indications. Gabapentin and pregabalin accounted for 40.8% and 19.6% of AED prescriptions in the comparator group respectively.

### 3.2 Comorbidities

Comorbidities are shown in **Table 2**. Compared to controls, PWE were significantly more likely to have intellectual disability, transient ischemic attack (TIA), intracranial hemorrhage, stroke, intracerebral abscess, other cerebrovascular disease, and traumatic brain injury as neurological comorbidities (these occurred <u>after</u> epilepsy diagnosis as all those with prior history of stroke/TIA were excluded). Hypertension, heart failure, coronary artery disease, valvular heart disease, and congenital heart disease were all more prevalent in PWE. Arrhythmias including atrial fibrillation/flutter (12.2% vs 6.9%; p<0.01), bradyarrhythmias (7.7% vs 3.5%; p<0.01), conduction defects (6.1% vs 2.6%; p<0.01), and ventricular arrhythmias (2.3% vs 1.0%; p<0.01), as well as cardiac implantable electric devices (4.6% vs 2.0%; p<0.01) were all more common in PWE compared to comparator group. (**Table 2**). PWE had higher resting heart rate than those without (70.84 vs 69.58; p=<0.001) and a greater proportion of PWE had resting heart rate > 100 BPM (1.8% vs 1.3%; p=0.001) (**Table 2**).

**Table 2.** Comorbidities and medications in epilepsy and comparator cohorts.

|  | **PWE** | **Comparator Group** | **P value** |
| --- | --- | --- | --- |
| N | 7,220 | 494,676 |  |
| Neurological comorbidities |  |  |  |
| Intellectual disability | 42 (0.6) | 139 (0.0) | <0.001 |
| Intracerebral abscess | 39 (0.5) | 119 (0.0) | <0.001 |
| TIA | 257 (3.6) | 6,273 (1.3) | <0.001 |
| Intracranial hemorrhage | 369 (5.1) | 3,868 (0.8) | <0.001 |
| Stroke* | 471 (6.5) | 13,340 (2.7) | <0.001 |
| Other cerebrovascular disease | 654 (9.1) | 9,766 (2.0) | <0.001 |
| Traumatic brain injury | 223 (3.1) | 2,515 (0.5) | <0.001 |
| Comorbidities |  |  |  |
| Diabetes | 994 (13.8) | 44,323 (13.8) | <0.001 |
| Valvular heart disease | 569 (7.9) | 20,004 (4.0) | <0.001 |
| Hypertension | 3,456 (47.9) | 183,547 (37.1) | <0.001 |
| Coronary artery disease | 803 (11.1) | 34,690 (7.0) | <0.001 |
| Atrial fibrillation/atrial flutter | 883 (12.2) | 34,007 (6.9) | <0.001 |
| Bradyarrhythmia | 555 (7.7) | 17,071 (3.5) | <0.001 |
| Conduction defect | 440 (6.1) | 12,646 (2.6) | <0.001 |
| CIED | 330 (4.6) | 9,964 (2.0) | <0.001 |
| Ventricular arrhythmia | 164 (2.3) | 4,893 (1.0) | <0.001 |
| Heart failure | 481 (6.7) | 15,149 (3.1) | <0.001 |
| Congenital heart disease | 60 (0.8) | 1,923 (0.4) | <0.001 |
| Medications |  |  | <0.001 |
| Beta blocker | 711 (9.8) | 33,518 (6.8) |  |
| Anti-arrhythmic | 138 (1.9) | 7,359 (1.5) |  |
| Mineralocorticoid antagonist | 31 (0.4) | 1,102 (0.2) |  |
| ACEi/ARB | 1,237 (17.1) | 72,708 (14.7) |  |
| AED | 3,514 (48.7) | 5,802 (1.2) |  |

\*Post epilepsy diagnosis; TIA = transient ischemic attack; CIED = cardiac implantable electronic device (including single- and dual-chamber permanent pacemaker, implantable cardioverter defibrillator, cardiac resynchronization therapy); ACEi = angiotensin-converting enzyme inhibitor; ARB = angiotensin II receptor blocker; AED = anti-epileptic drug. For continuous variables standard deviations are enclosed within brackets; for categorical variables percentages are enclosed.

### 3.3 Factors influencing mortality

There were 6,171,803 person-years of follow-up (mean 12.30 years, SD 1.74). Only 1,298 (0.26%) of participants were lost to follow-up (1,284 controls and 14 PWE). PWE died at a younger age [68.1 vs 69.8 years; p<0.001]. This is equivalent to 2,230 person-years of life lost. (**Table 1**). Kaplan-Meier plots for all-cause and sudden death-specific mortality are shown in **Figure 2**. In univariable Cox analysis, survival was significantly worse in PWE (HR 3.37[95% CI, 3.18-3.56]) (**Figure 2A**). Survival remained significantly worse in PWE even after adjusting for male sex, intellectual disability, taking more than 2 AEDs, and generalized epilepsy (HR 3.19 [95% CI, 3.01-3.39]) in multivariable Cox proportional hazard regression. PWE were also significantly more likely to suffer sudden death events (**Figure 2B**) (sudden death-specific HR 6.65 (95% CI, 4.53-9.77) and the subdistribution HR 5.06 (95% CI, 3.46-7.40)).

**Figure 2.**
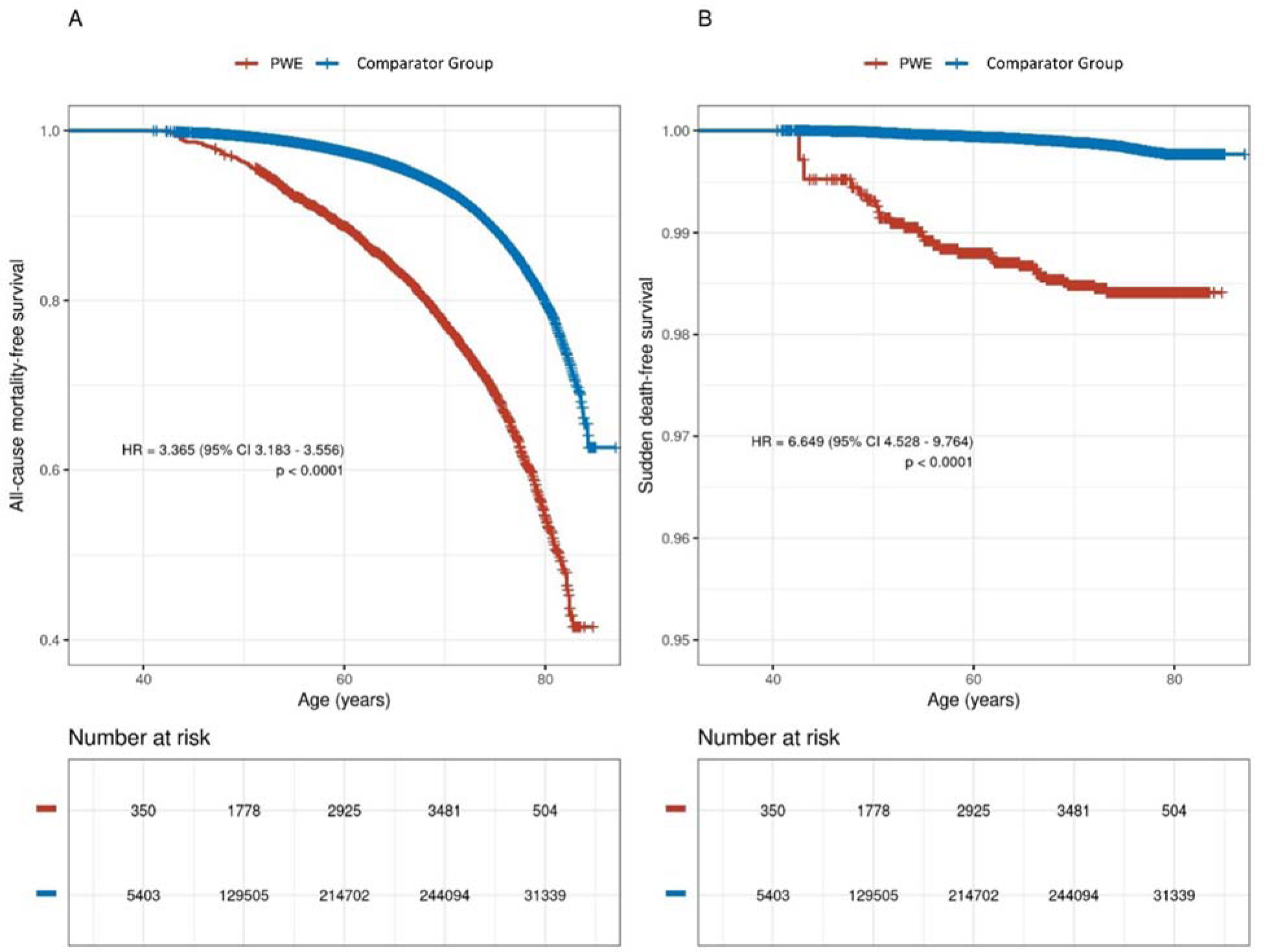
Kaplan-Meier curves for all-cause mortality-free survival (**A**) and sudden death-free survival (**B**) in epilepsy and comparator group population. Hazard ratios reflect Cox proportional hazards regressions modelling the cohorts as left-truncated and right-censored, with epilepsy modelled as a time-dependent covariate and additionally controlled for sex.

**Figure 3.**
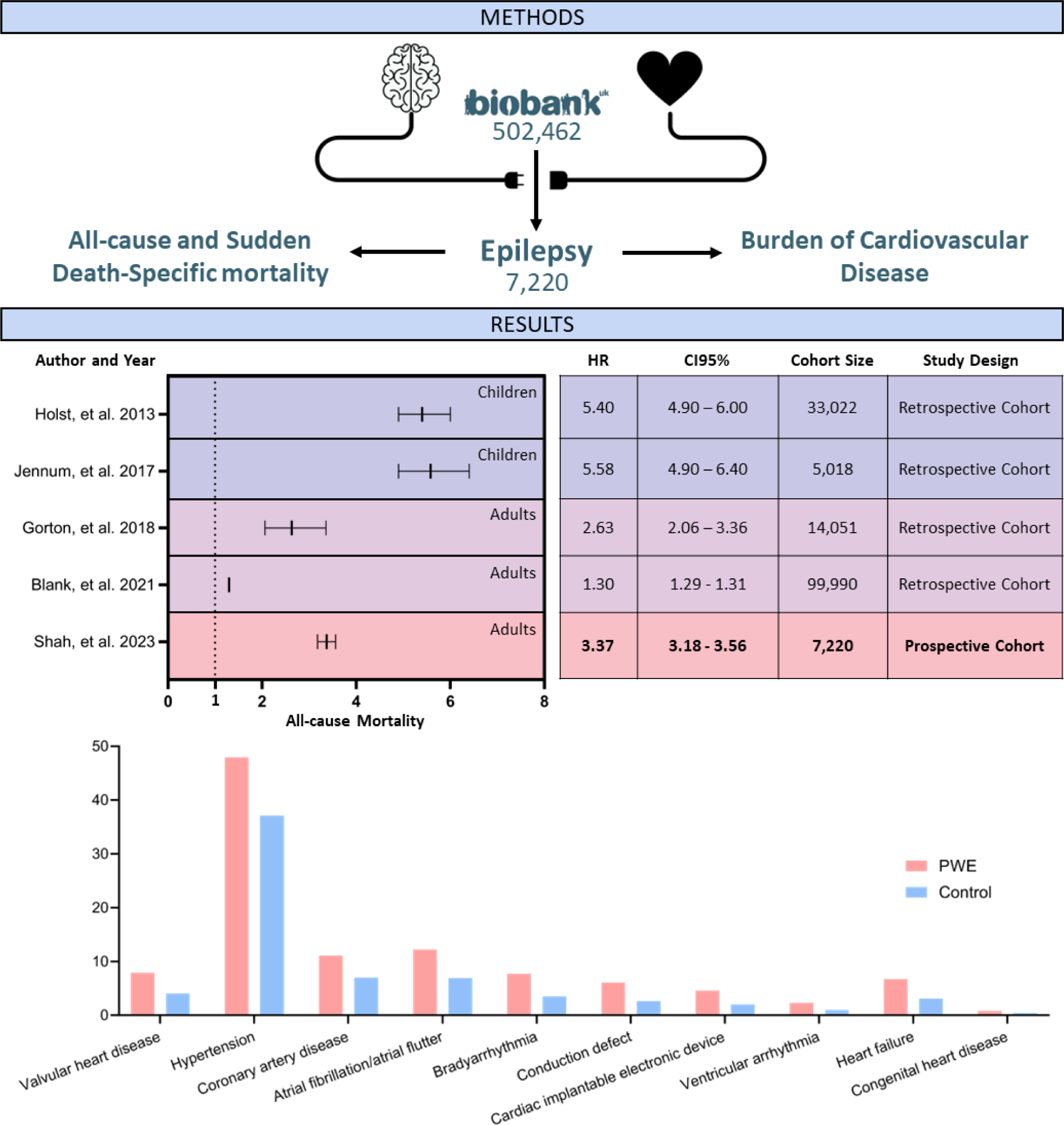
Central illustration

Univariable and multivariable Cox proportional hazard regression showed male sex and intellectual disability were both independently associated with significant worsening of all-cause survival in comparator group (**eTable4**). In PWE, male sex and generalized epilepsy had influence on mortality in univariable and multivariable Cox regression (**eTable 4**). Taking ≥3 AEDs was not associated with higher all-cause mortality in PWE (**eTable 4**). The effect of male sex on all-cause mortality was smaller in PWE compared with the entire cohort (p = 0.0086).

We also analyzed the effect of obesity on survival. BMI of more than 30 kg/m^2^ was associated with higher mortality (HR 1.41 [95% CI 1.37-1.44]) in the overall population; however, there was no effect of BMI of more than 30 kg/m^2^ in PWE (**eTable 5**). Higher resting heart rate was associated with increased mortality in both PWE and the overall population (HR 1.02 (95% CI 1.01-1.02) (**eTable 6**).

## 4. Discussion

To our knowledge, this is the largest prospective cohort of PWE with the longest person-years of follow-up investigating all-cause mortality and sudden death. PWE have markedly higher prevalence of CVD than controls. We have found that middle-aged PWE have a 3-times higher all-cause mortality and 6-fold higher risk of sudden death, with younger age at death. PWE had higher resting heart rates and this was also associated with mortality in both PWE and controls, though worse in PWE.

We have found markedly higher prevalence of CVD in PWE than controls. The BMI difference was marginal and is insufficient to account for higher hypertension. Congenital heart disease and valvular heart disease may be explained by syndromic conditions with congenital heart disease and epilepsy. However, the higher burden of coronary artery disease and heart failure warrant further investigation. Increased rates of arrhythmia and conduction disease in PWE compared to the comparator group, and therefore increased rates of cardiovascular disease may be contributing to the excess mortality among PWE. **A recent term used to describe this is the “epileptic heart” where** "a heart and coronary vasculature damaged by chronic epilepsy as a result of repeated surges in catecholamines and hypoxemia leading to electrical and mechanical dysfunction." When arrhythmias are observed in SUDEP, the majority are bradyarrhythmias rather than tachy-arrhythmias, with reported cases of syncope with temporal lobe epilepsy resolved with the implantation of a pacemaker.^11, 23, 24^ This is of importance given most focus of SUDEP research and policymakers is on those under 40 years of age.^10, 25^ However, it would be illogical to assume that SUDEP does not occur <u>after</u> the age of 40 years. The findings from this prospective study (vs published <u>retrospective</u> analyses) support increased risk of arrhythmias and also structural disease including CAD, valvular heart disease and heart failure. Even after adjustment and matching by age and sex, and removing those with disease at enrollment, this persisted. While this does not fulfill criteria for SUDEP, because of lack of autopsy data, the increased all-cause mortality and sudden unexpected death-specific mortality should not be ignored. Investigating this observation is warranted, as it may explain a subset of sudden deaths in epilepsy and may challenge the current definitions of SUDEP.

Our data reproduce and support earlier findings of increased risk of both all-cause and sudden death-specific mortality in PWE.^2, 4, 14–17, 26–29^ These studies have been critiqued for containing high proportions of persons with <u>refractory</u> epilepsy reflecting selection and tertiary center referral bias, or studying cohorts admitted to epilepsy monitoring units.^14, 15, 28^ Our study analyzed outcomes in 7,220 PWE and to date is the largest reported cohort.^17, 26^ Additionally, the PWE population reported here was derived from a UKBB population and is more reflective of a general population, although there are limitations with non-White populations and healthy volunteer bias. All outcome data in the UKBB were collected prospectively with very little loss of follow-up, reducing the risk of bias and increasing confidence in the robustness of the data. Thus, these observations are likely a minimum estimate of CVD burden, all-cause and sudden death-specific mortality.

We aimed to assess the effect of factors previously identified as being associated with epilepsy mortality. Most of these factors were determined in SUDEP cases.^7–9^ We had insufficient SUDEP numbers to assess whether these predictors had validity in this cohort of PWE and, therefore, we assessed their impact on overall mortality instead. With the available data within the UKBB, we were able to test the following predictors: male sex, intellectual disability, generalized epilepsy, and polytherapy.^7, 9^ We found that males had higher mortality, similar to previous studies;^7^ however this effect size was smaller in the PWE cohort compared to the comparator group. The importance of intellectual disability is disputed, with some studies finding evidence supporting this association^9^ and some reporting no effect.^7, 8^ The study highlighting the importance of intellectual disability included 20 SUDEP cases^9^ compared to more than 200 cases in the two refuting this.^7, 8^ We found no effect of intellectual disability on outcome in our epilepsy cohort despite an effect in comparator group. Polypharmacy has been identified as a risk factor for SUDEP;^7–9^ this may, in fact, reflect more severe epilepsy, which is difficult to control, rather than the effects of polypharmacy directly. However, we did not find any effect of polypharmacy on overall mortality rates in PWE. This may be explained by polytherapy being a risk factor for SUDEP-specific mortality only, and not for all-cause mortality. We also found that generalized epilepsy is associated with increased risk of overall mortality similar to previous studies. ^7–9^

Higher all-cause and sudden death specific mortality, which has been previously noted in those younger than 40 years of age, also manifests in middle-aged persons with epilepsy. We have found large, clinically significant effect sizes with a 3-fold higher risk of all-cause mortality and 6-fold higher risk of sudden death specific mortality. We have found that resting heart rate and generalized epilepsy is associated with greater risk of mortality. As resting heart rate is a known risk factor for sudden cardiac death,^30, 31^ SUDEP may share similar disease mechanisms to sudden cardiac death. Overall, these findings highlight an important area of future research into mortality mechanisms so that management and outcomes of these individuals can be improved.

Strengths of our study include (1) large sample size of cases and controls; (2) extremely low number of participants lost to follow-up; (3) a long duration of follow-up; (4) reliable and accurate mortality data; and (5) a population aged between 40-69 years allowing us to investigate this area of conflicting evidence. Previous studies have suggested the greatest mortality rates are in younger patients with epilepsy,^17^ with one study finding the greatest excess mortality in those aged 31-40 years.^27^ Older PWE are assumed to be at lower risk of mortality than those aged < 40 years. The hazard ratios and narrow confidence intervals reported in this study challenge these assertions. The main limitation of the study is that the UKBB population is healthier than the general UK population, displaying healthy volunteer bias. This considered, the risk of mortality and sudden death may be even higher than we report. Secondly, as the UKBB only enrolls 40-69 years, our study may be affected by survivor bias where individuals with high-risk epilepsy may have died before they could be recruited into the study. Thirdly, the diagnosis was based on linked hospital diagnostic codes for epilepsy and death certificates to determine cause of death. This is liable to inaccuracy of the epilepsy diagnosis itself, categorization to epilepsy subgroups, and identifying cases of sudden death. Another limitation is lack of data on subtype of epilepsy, nocturnal GTCS, frequency of seizures, and sleep disordered breathing. Considering the above, the burden of CVD is a minimum estimate and in reality could be much higher. Similarly, the all-cause mortality and sudden death risk, are likely minimal estimates.

## 5. Conclusions

In this large population-based cohort study, we have found that middle-aged PWE have a high prevalence of CVD, increased risk of both all-cause and sudden death-specific mortality. PWE had higher resting heart rates and this was associated with increased mortality. PWE with a subtype of generalized epilepsy had increased mortality. In PWE, intellectual disability and polypharmacy was not associated with increased mortality. Further work is required to elucidate mechanisms underlying both all-cause mortality and sudden death risk and identifying biomarkers of prognosis in PWE.

## Data Availability

Data is available per UK Biobank policy.

## Non-standard Abbreviations and Keywords

AED: Anti-epileptic drug
CVD: Cardiovascular disease
PWE: Persons with epilepsy
UKBB: UK Biobank

## Acknowledgments

None.

